# SARS-CoV-2 vaccination predicts COVID-19 progression and outcomes in hospitalized patients

**DOI:** 10.1101/2021.11.21.21266633

**Authors:** Alessandro Padovani, Viviana Cristillo, Daniela Tomasoni, Stefano Gipponi, Andrea Pilotto

**Affiliations:** Department of Clinical and Experimental Sciences, University of Brescia, Italy; Multidisciplinary COVID-Unit, ASST Spedali Civili Hospital, Brescia, Italy; Department of Medical and Surgical Specialities, Radiological Sciences and Public Health, University of Brescia, Brescia, Italy

**Keywords:** COVID-19, Vaccination, disease progression, lethality, death

## Abstract

**Background:** SARS-CoV-2 vaccination might impact on clinical progression of cases with breakthrough COVID-19 disease.

**Objective:** to evaluate the impact of SARS-CoV-2 vaccination on disease progression in COVID-19 hospitalized patient

**Methods and Findings:** Two-hundred eighty-four consecutive COVID-19 hospitalized patients, including 50 vaccinated cases entered the study. Compared to unvaccinated cases, vaccinated patients were older, exhibited more comorbidities and did not differ for COVID-19 severity at admission. During hospitalisation, unvaccinated patients showed worse disease progression, including higher need of oxygen and higher risk of death compared to vaccinated patients (OR 3.3; 1.05-10.7 95% CI in the whole cohort and OR 54.8; 3.5-852 in the ventilated cases).

**Discussion:** These findings argue for an important reduction in severity among vaccine breakthrough infection compared to unvaccinated cases in COVID-19 disease.

## Background

Vaccination against SARS-CoV-2 consistently exhibited high efficacy in reducing the incidence of COVID-19 either in trials [1] or in real-world settings [2,3]. Nevertheless, COVID-19 disease has been reported in patients who underwent SARS-Cov-2 vaccination [4]. A recent study has demonstrated that mRNA vaccination was significantly less likely among hospitalized patients with severe COVID-19 [5].

## Objective

In this observational study, we aimed at evaluating the impact of vaccination on disease progression, by controlling for premorbid conditions and severity at admission.

## Methods and Findings

The study included all consecutive patients hospitalized for SARS-CoV-2 infection at the multidisciplinary COVID-Unit of the ASST Spedali Civili Hospital, Brescia, from March 1 to October 15, 2021. At admission pre-morbid comorbidities and general health conditions were assessed by the Cumulative Illness Rating Scale (CIRS) and the modified Rankin scale (mR) while COVID-19 clinical severity was measured by the Brescia-COVID Respiratory Severity Scale (BCRSS) and the National Early Warning Score (NEWS)2. Outcome measures were collected during hospitalization and included need of steroid, remdesivir or oxygen treatment, invasive ventilation and death. The World Health Organisation COVID-19 Clinical progression scale was used as general progression measure, as it ranges from infected but asymptomatic (level 1) to death (level 9). The study received approval from local ethics committee of the ASST Spedali Civili Hospital, Brescia (NP 4067, approved 08.05. 2020). Differences in demographics characteristics and premorbid conditions were evaluated in vaccinated and non-vaccinated subjects. A linear and logistic regression model was implemented in order to evaluate the differences in clinical outcomes between vaccinated and unvaccinated subjects adjusted for the effect of premorbid conditions and admission severity.

Two-hundred eighty-four consecutive COVID-19 hospitalized patients entered the study. Out of the total sample, 50 patients (17.6%) received Covid-19 vaccination, namely 18 and 32 cases immunized with a single and full dose, respectively. Almost all vaccinated patients (84%) received a messenger RNA vaccine (BNT162b2 Pfizer–BioNTech and mRNA-1273 Moderna), while 8 patients received a viral vectored coronavirus vaccine (ChAdOx1 nCoV-19 Oxford–AstraZeneca and Ad26.COV2.S Janssen-Johnson and Johnson).

Vaccinated patients were significantly older (73.9±13.7 vs. 65.3±17.5, p=0.001), showed more comorbidities (p=0.013) but they did not differ from unvaccinated patients for COVID-19 severity at admission (Table 1). During hospitalisation, unvaccinated patients showed a worse progression (WHO Clinical Progression Scale 5.0±1.4 vs 5.3±1.6; p<.03), higher need of oxygen (p=0.048) and steroid (p=0.002). Multivariate regression analyses adjusted for age and comorbidities showed a higher risk of death in unvaccinated cases in the whole cohort (OR 3.3; 1.05-10.7 95% CI) and in ventilated patients (OR 54.8; 3.5-852). Similar findings were obtained in older patients ≥65 years old (n=162): unvaccinated subjects showed a higher death risk (OR 3.2; 1.02-10.5; p<.0.04) which increased in ventilated subjects (OR 62.6; 2.9-1327; p<.008).

**Table 1.** Demographic, clinical, laboratory characteristics of Covid-19 patients according to the SARS-CoV-2 vaccination. \**p* values were calculated by t-test or Fisher’s exact test, as appropriate. #p-value corrected for the effect of age and CIRS by multivariate regression analyses. Abbreviations: BCRSS, Brescia-COVID Respiratory Severity Scale; CIRS, Cumulative Illness Rating Scale; News2, National Early Warning Score 2.

|  | Unvaccinated<br>(n=234) | Vaccinated<br>(n=50) | * *p* Value |
| --- | --- | --- | --- |
| **Clinical and demographics features** |  |  |  |
| Age, years | 65.3±17.5 | 73.9±13.7 | **0.001** |
| Sex, female | 92 (39.3%) | 121(42.0%) | 0.752 |
| Days of hospitalization, n | 13.8±12.6 | 12.9±9.9 | 0.801 |
| Modified Rankin Scale premorbid | 1.41±1.2 | 1.67±1.3 | 0.050 |
| CIRS pre-admission total value | 19.6±4.2 | 21.6±4.1 | **0.005** |
| BCRSS at admission | 0.69±0.9 | 0.66±0.9 | 0.514 |
| News2 at admission | 2.39±2.3 | 2.10±1.8 | 0.313 |
| **Outcomes measures** |  |  |  |
| WHO COVID-19 Clinical progression scale | 5.3±1.6 | 5.0±1.4 | **0.030#** |
| Need of Oxygen treatment | 141 (60.3%) | 26 (52.0%) | **0.048#** |
| Steroid treatment | 148 (63.2%) | 20 (40.0%) | **0.002#** |
| Remdesivir treatment | 22 (9.4%) | 6 (12%) | 0.42# |
| Need of ventilation | 42 (17.9%) | 10 (20.0%) | 0.41# |
| Death | 33 (14.1%) | 4 (8%) | **0.041#** |

## Discussion

These findings are in agreement with epidemiological data claiming for a reduction of raw mortality in vaccinated subjects, independently from the development of the disease [1-5]. In addition, our study showed that vaccination has a protective effect on clinical course by reducing the progression and lethality, even in the elderly with elevated multimorbidity. In particular, the positive effect on lethality was dramatic among the most severe cases, as the risk of death increased from 3.3 fold in the whole cohort to more than 50 fold in ventilated subjects.

As main limitation of the present study, we did not evaluate the IgG antibodies title data for immunized patients, which might be associate with different degree of systemic and respiratory immune response to SASR-CoV-2 infection. We also acknowledged that the statistical models were limited by the small sample size and that multivariate analyses including prognostic factors or propensity algorithms need to be implemented in on-going multi-center prospective studies. Limitations notwithstanding, our findings argue for a beneficial role of vaccination beyond the prevention of the disease, that dramatically impact on clinical progression and outcomes in COVID-19 disease.

## Data Availability

l data produced in the present study are available upon reasonable request to the authors

